# Little Evidence of Modified Genetic Effect of rs16969968 on Heavy Smoking Based on Age of Onset of Smoking

**DOI:** 10.1101/2020.04.22.20071407

**Authors:** Christine Adjangba, Richard Border, Pamela N. Romero Villela, Marissa A. Ehringer, Luke M. Evans

**Affiliations:** Institute for Behavioral Genetics, University of Colorado Boulder; Department of Applied Mathematics, University of Colorado Boulder; Department of Psychology & Neuroscience, University of Colorado Boulder; Department of Integrative Physiology, University of Colorado Boulder; Department of Ecology & Evolutionary Biology, University of Colorado Boulder

## Abstract

Tobacco smoking is the leading cause of preventable death globally. Smoking quantity, measured in cigarettes per day (CPD), is influenced both by the age of onset of regular smoking (AOS) and by genetic factors, including a strong effect of the non-synonymous single nucleotide polymorphism rs16969968. A previous study by Hartz et al. reported an interaction between these two factors, whereby rs16969968 risk allele carriers who started smoking earlier showed increased risk for heavy smoking compared to those who started later. This finding has yet to be replicated in a large, independent sample. We performed a preregistered, direct replication attempt of the rs16969968×AOS interaction on smoking quantity in 128,383 unrelated individuals from the UK Biobank, meta-analyzed across ancestry groups. We fit statistical association models mirroring the original publication as well as formal interaction tests on multiple phenotypic and analytical scales. We replicated the main effects of rs16969968 and AOS on CPD but failed to replicate the interaction using previous methods. Nominal significance of the rs16969968×AOS interaction term depended strongly on the scale of analysis and the particular phenotype, as did associations stratified by early/late AOS. No interaction tests passed genome-wide correction (α=5e-8), and all estimated interaction effect sizes were much smaller in magnitude than previous estimates. We failed to replicate the strong rs16969968×AOS interaction effect previously reported. If such gene-moderator interactions influence complex traits, they likely depend on scale of measurement, and current biobanks lack the power to detect significant genome-wide associations given the minute effect sizes expected.

**IMPLICATIONS:** We failed to replicate the strong rs16969968×AOS interaction effect on smoking quantity previously reported. If such gene-moderator interactions influence complex traits, current biobanks lack the power to detect significant genome-wide associations given the minute effect sizes expected. Furthermore, many potential interaction effects are likely to depend on the scale of measurement employed.

## INTRODUCTION

Approximately 20% of deaths every year in the United States can be attributed to cigarette smoking, and smokers have life expectancies at least 10 years shorter than nonsmokers^1^. Furthermore, the rise in use among adolescents of various electronic cigarettes has emerged as a potentially dangerous trend about which little is known regarding long-term health and addiction consequences^2^. There is strong evidence from adoption, family, and twin studies that both genetic and environmental factors contribute to risk for smoking behaviors, with heritability estimates for nicotine dependence, ever becoming a regular smoker, and smoking quantity ranging between 33% and 71%^3-7.^ Recently, genome-wide association studies (GWAS) have identified common variants associated with smoking^8-13^. In particular, the nicotinic acetylcholine receptor subunit genes *CHRNA5-CHRNA3-CHRNB4* on chromosome 15 have been implicated by well-powered GWAS of smoking behaviors^12,14,15^. Within *CHRNA5*, which codes for the α5 receptor subunit, the nonsynonymous G/A single nucleotide polymorphism (SNP) rs16969968 has been replicated through both large-scale GWAS^8-13,16^ and functional assays^17-20^ to influence smoking quantity, as measured by the number of cigarettes smoked per day, and nicotine dependence. The rs16969968-A risk allele has the largest estimated allelic effect on smoking quantity known to date^12^. While GWAS have identified many additional smoking-associated variants, rs16969968 remains a focus of individual functional studies and genetic epidemiological studies, with 292 publications reporting analyses of rs16969968 indexed by dbSNP (https://www.ncbi.nlm.nih.gov/snp/rs16969968#publications) and 454 publications (198 within the last five years) listed on LitVar (www.ncbi.nlm.nih.gov/CBBresearch/Lu/Demo/LitVar), at the time of this writing.

In addition to genetic risk factors for heavy smoking, earlier age at onset of regular smoking (AOS) is well-known to predict risk for later heavy use and nicotine dependence^21,22^. In light of previous findings, Hartz et al.^13^ conducted a meta-analysis of 33,348 individuals across 43 European and American data sets to test whether genetic vulnerability to heavy smoking and nicotine dependence at rs16969968 depends on AOS, and found a strong, significant interaction between early AOS and the rs16969968-A allele on heavy smoking (OR=1.16). Additional studies^23,24^ have focused on rs16969968 interactions with other variables, highlighting the continued interest in rs16969968 interactions on behavior. Notably, these include those evaluating rs16969968×age of nicotine exposure^20,25^, and include a report that early intervention to prevent adolescent smoking reduces the genetic risk of rs16969968 for heavy smoking later in life, a gene-by-intervention interaction^26^. The original finding of rs16969968×AOS has been referenced in reviews citing the need for evaluation of gene-by-environment (G×E) interaction effects on nicotine dependence ^27^, and suggesting that direct replication of methods is needed to rigorously evaluate G×E interactions on smoking^28^. However, despite the large rs16969968×AOS interaction effect size originally reported, animal model evidence to support the plausibility of such an interaction^20^, and continued interest in rs16969968, we are aware of no large-scale replication attempt in an independent sample. Here, we assessed whether there is an rs16969968×age of onset of smoking interaction in a well-powered (Fig. S1), independent sample, in an attempt to directly replicate the original findings.

## METHODS

We preregistered our analyses through the Open Science Framework (osf.io/ynh2j) after we had obtained the UK Biobank data, but before we analyzed CPD or AOS.

### Study Population

We used the UK Biobank, a large sample with rich phenotype and genome-wide genotype data^29^. We included all participants with available genomic data who had reported CPD and AOS data. The participants were either current or former smokers aged 40 years or older. To avoid confounding influences of population stratification^30^, we initially, and following our preregistration, performed analyses using only individuals of European (EUR) ancestry, the largest subsample within the UK Biobank, identified by those whose first scores on the first four principal components (PCs; UK Biobank data field ID 22009) fell within the range of the UK Biobank identified individuals of European ancestry (field 22006). Following this, we expanded our analysis to include all available individuals within the UK Biobank. We identified relatively genetically homogeneous groups of individuals within the UK Biobank after excluding the EUR-ancestry individuals noted above using K-means clustering, from K=2-10, applied to the first 10 PC axes (data field ID 22009). The percent variance explained plateaued at K=10 clusters (Fig. S2). All analyses were subsequently performed within these 11 genetic clusters (EUR-ancestry + K=10 clusters). We note that the purpose of this clustering was solely to identify relatively genetically homogeneous groups of individuals within which to perform association analyses, and not to make population genetic inferences.

We only included unrelated individuals in our primary analyses to avoid possible confounding due to shared environmental factors. Relatedness was estimated within each genetic cluster using MAF- and LD-pruned array markers (plink2^31^ command: --maf 0.01 --hwe 1e-8 --indep-pairwise 50 5 0.2) after excluding those individuals with self-report and genetic sex mismatch (fields 31 and 22001), those with unusually high inbreeding coefficients (|F_het_| >0.2), and those identified by the UK Biobank and Affymetrix as having poor-quality genomic data (fields 220010 and 22051). Unrelated individuals (estimated relatedness<0.05) were identified with GCTA^32^ v1.91.3 within each cluster. After removing individuals with missing phenotype and covariate data (see below), a total of 128,383 unrelated individuals across all genetic clusters were included.

### Variables

Smoking quantity, as measured by CPD, was the primary dependent variable in analyses. Data on CPD (fields 2887, 3456, and 6183) were obtained from current or former smokers by asking the question “About how many cigarettes do/did you smoke on average each day?” These data were highly skewed; therefore, we also analyzed log_10_-transformed CPD (Fig. S3). Because of observed evidence of scale dependence^33^ (see results below), we also analyzed heavy/light CPD on an additive scale. These two additional procedures were the only deviations from our preregistered analyses. Final analyses considered untransformed CPD, log_10_(CPD), heavy/light (analyzed on both multiplicative, i.e., logistic, and additive scales), and binned encodings. The dichotomous encoding defined smoking quantity as light smoking (CPD ≤ 10) versus heavy smoking (CPD > 20), mirroring the definition used by Hartz et al. The binned encoding defined smoking quantity as a linear variable consisting of 0 (CPD ≤ 10), 1 (11-20 CPD), 2 (21-30 CPD), or 3 (CPD > 30), also matching their secondary analysis.

Age of onset of regular smoking (AOS) was determined from fields 3426 and 2867, where participants were asked “How old were you when you first started smoking on most days?” AOS was analyzed based on a dichotomous encoding, a binned encoding, and the raw AOS data, again replicating the methods of Hartz et al.^13^. The dichotomous encoding defined early as AOS ≤ 16 years and late as AOS > 16 years. The binned encodings were 0 (AOS ≤ 15 years), 1 (AOS = 16 years), 2 (17-18 years AOS), or 3 (AOS > 18 years). We note that, matching Hartz et al., the median AOS was 16 in the UK Biobank (Fig. S3), making a reasonable and comparable age at which to separate early vs. late initiating smokers.

Covariates included were sex (field 31), age at time of assessment (field 21003), age^2^, Townsend Deprivation Index (field 21003), educational attainment (“qualification”, categorical, field 6138), genotyping batch (field 22000), assessment center (field 54), and the first 10 genetic principal components as estimated with flashpca^34^ applied to the MAF- and LD-pruned SNPs as described above. High collinearity of covariates within this sample resulted in a rank-deficient design matrix, which we addressed by performing a principal components analysis of the *c*=141 fixed effects using the prcomp function in R v3.2.2^35^. We then estimated the rank of the resulting eigenvector matrix (rank *r* < *c*) using the *matrix* R package^36^ and included the first *r*=140 principal components as covariates in all analyses.

### Statistical Analyses

All statistical analyses were performed within each genetic ancestry cluster separately. For dichotomized light/heavy CPD, we performed logistic regression using *glm* (family=‘binomial’) in R^35^ to assess the multiplicative scale interaction. The model included the rs16969968 genotype (coded as 0, 1, or 2), AOS, and rs16969968×AOS. All genotype×covariate and AOS×covariate interactions were included within the models to appropriately control for confounding^37^. For continuous variables (raw, binned, and log-transformed CPD) and the additive scale interaction model of the dichotomous heavy/light phenotype, we tested the same model using linear regression with the R *lm* function. Because many of the non-EUR-ancestry clusters had relatively few unrelated individuals within them, including all 140 covariates and their interactions resulted in a model that could not be fitted. We therefore reduced the number of covariates to be the scores from the first five PC scores of the covariate design matrix for the K=10 non-EUR-ancestry clusters.

The above model varied from that tested by Hartz et al., who tested rs16969968 effects on smoking phenotypes stratified by AOS (early versus late), using logistic regression (i.e., multiplicative scale). To recapitulate their methods, we performed secondary association tests of rs16969968 stratified by early versus late AOS using BOLT-LMM v2.3.2^38^, with 339,444 genome-wide SNPs (quality control as described above, but without LD-pruning) to control for cryptic relatedness. All covariates were included in the BOLT-LMM models, excluding interaction terms. Because BOLT-LMM is not recommended for samples of less than 5,000 (see documentation from ref. ^38^), we used GCTA leave-one-chromosome-out (--mlma-loco) approach^39^ for the non-EUR-ancestry genetic clusters. Finally, to directly replicate previous methods, we performed AOS-stratified logistic regression of heavy/light CPD using only rs16969968 and sex as independent variables.

We meta-analyzed the results using the inverse variance weighting approach in METAL^40^. We report meta-analyzed results below, and all cluster-specific results in the Supplementary Material.

We also performed several power analyses, to determine the power to detect the previously reported effect size^13^, as well as to determine the sample size needed to achieve 80% power at specified effect sizes and α. To estimate the power to detect the previously reported effect size in the UK Biobank sample under a multiplicative scale interaction model, we simulated 61,077 diploid genotypes and early/late AOS in R, with linear predictors simulated using the previously reported main effect sizes as,

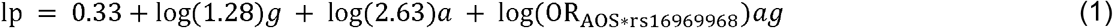

where genotypes, *g*, were simulated from a binomial distribution with MAF=0.34, the observed frequency of the A allele in the UK Biobank, early versus late AOS status, *a*, was randomly assigned to individuals. We varied the interaction effect size, log(ORAOS*rs16969968) between 0.005 and 0.4, reflecting a range of plausible effect sizes and encompassing the previously reported interaction effect (OR=1.16). Binary phenotypes, y, were then simulated in R as,

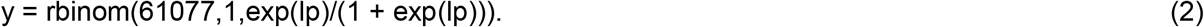

For each simulated interaction effect size, we performed 1,000 replicate simulations, estimating the interaction effect using logistic regression as above, and recorded the number of observations with an interaction *p*-value below either nominal significance, α=0.05, or genome-wide significance, α=5e-8. We performed similar simulations with the main AOS and rs16969968 effect sizes estimated within the UK Biobank (see below). We varied the sample size from 1e3 to 2e6, varying interaction effect size (previously reported ORAOS*rs16969968=1.16 versus our meta-analyzed estimate ORAOS*rs16969968=1.004), and nominal versus genome-wide significance thresholds (α=0.05 versus 5e-8, respectively).

## RESULTS

We observed significant main effects of the rs16969968 A allele and AOS on CPD (Figures 1A, 1B, S4; Tables 1, S1-2). When estimated as predictors of heavy vs. light smoker status, the meta-analyzed estimated genetic effect, ORrs16969968=1.12 (*p*=4.8e-28), was similar to but lower than the previous estimate^13^ of 1.28. The effect of early AOS, ORAOS=1.19 (*p*=3.6e-45), was less than previously reported^13^ (ORAOS=2.63). However, both main effects were significantly associated with CPD in the expected direction, regardless of the CPD or AOS encoding, and represent strong evidence that both the rs16969968 A allele and early AOS are positively associated with heavier smoking, replicating previous findings. We note that, like previous findings^12,13^, rs16969968 is not associated with AOS (Fig. S3D)

**Table 1.** Trans-ethnic meta-analysis estimated main and interaction effects (β) and standard errors (SE) for rs16969968, age of smoking initiation (AOS), and their interaction. Shown are estimates for each encoding of CPD and AOS.

| CPD Coding | N | AOS Coding | AOS |  |  | rs16969968_A |  |  | rs16969968 x AOS |  |  |
| --- | --- | --- | --- | --- | --- | --- | --- | --- | --- | --- | --- |
| | | | $\beta^a$ | SE | p | $\beta^a$ | SE | p | $\beta^a$ | SE | p |
| CPD (raw) | 128383 | raw | -0.296 | 1.15E-02 | 9.34E-147 | 1.271 | 2.17E-01 | 4.65E-09 | -0.019 | 1.24E-02 | 1.17E-01 |
|  |  | binned | -0.830 | 3.59E-02 | 3.89E-118 | 1.042 | 6.53E-02 | 2.57E-57 | -0.063 | 3.81E-02 | 1.01E-01 |
|  |  | Early/Late | 1.589 | 8.18E-02 | 4.99E-84 | 0.861 | 5.97E-02 | 3.41E-47 | 0.190 | 8.71E-02 | 2.87E-02 * |
| log<sub>10</sub>(CPD) | 128383 | raw | -0.008 | 2.88E-04 | 6.91E-159 | 0.026 | 5.36E-03 | 1.55E-06 | 0.000 | 3.07E-04 | 7.86E-01 |
|  |  | binned | -0.021 | 8.83E-04 | 1.80E-129 | 0.025 | 1.60E-03 | 4.22E-54 | 0.000 | 9.34E-04 | 8.19E-01 |
|  |  | Early/Late | 0.040 | 2.01E-03 | 1.95E-86 | 0.023 | 1.46E-03 | 3.40E-56 | 0.003 | 2.13E-03 | 2.12E-01 |
| binned | 128383 | raw | -0.023 | 9.39E-04 | 1.10E-130 | 0.102 | 1.77E-02 | 9.89E-09 | -0.001 | 1.01E-03 | 2.03E-01 |
|  |  | binned | -0.065 | 2.93E-03 | 3.07E-109 | 0.089 | 5.32E-03 | 4.94E-63 | -0.006 | 3.11E-03 | 6.59E-02 |
|  |  | Early/Late | 0.126 | 6.67E-03 | 2.55E-79 | 0.074 | 4.87E-03 | 1.26E-52 | 0.015 | 7.10E-03 | 3.79E-02 * |
| Heavy/Light | 61077 | raw | -0.018 | 1.41E-03 | 1.15E-38 | 0.116 | 3.94E-02 | 3.16E-03 | 0.000 | 2.16E-03 | 9.00E-01 |
|  |  | binned | -0.077 | 4.92E-03 | 1.23E-54 | 0.146 | 1.27E-02 | 2.78E-30 | -0.001 | 6.80E-03 | 8.30E-01 |
|  |  | Early/Late | 0.171 | 1.21E-02 | 3.65E-45 | 0.116 | 1.05E-02 | 4.82E-28 | 0.004 | 1.62E-02 | 8.22E-01 |
| Heavy/Light<br>(additive scale) | 61077 | raw | -0.013 | 7.19E-04 | 6.59E-76 | 0.094 | 1.43E-02 | 5.02E-11 | -0.002 | 8.15E-04 | 2.87E-02 * |
|  |  | binned | -0.044 | 2.23E-03 | 2.40E-88 | 0.070 | 4.24E-03 | 2.40E-61 | -0.005 | 2.45E-03 | 5.70E-02 * |
|  |  | Early/Late | 0.090 | 5.17E-03 | 4.34E-67 | 0.058 | 3.88E-03 | 7.84E-50 | 0.012 | 5.69E-03 | 3.41E-02 |

<sup>a</sup> $\beta$ refers to the regression slope. For CPD coded as heavy/light, $\exp(\beta)$ is the Odds Ratio (OR) when analyzed on the multiplicative scale.

**Figure 1.**
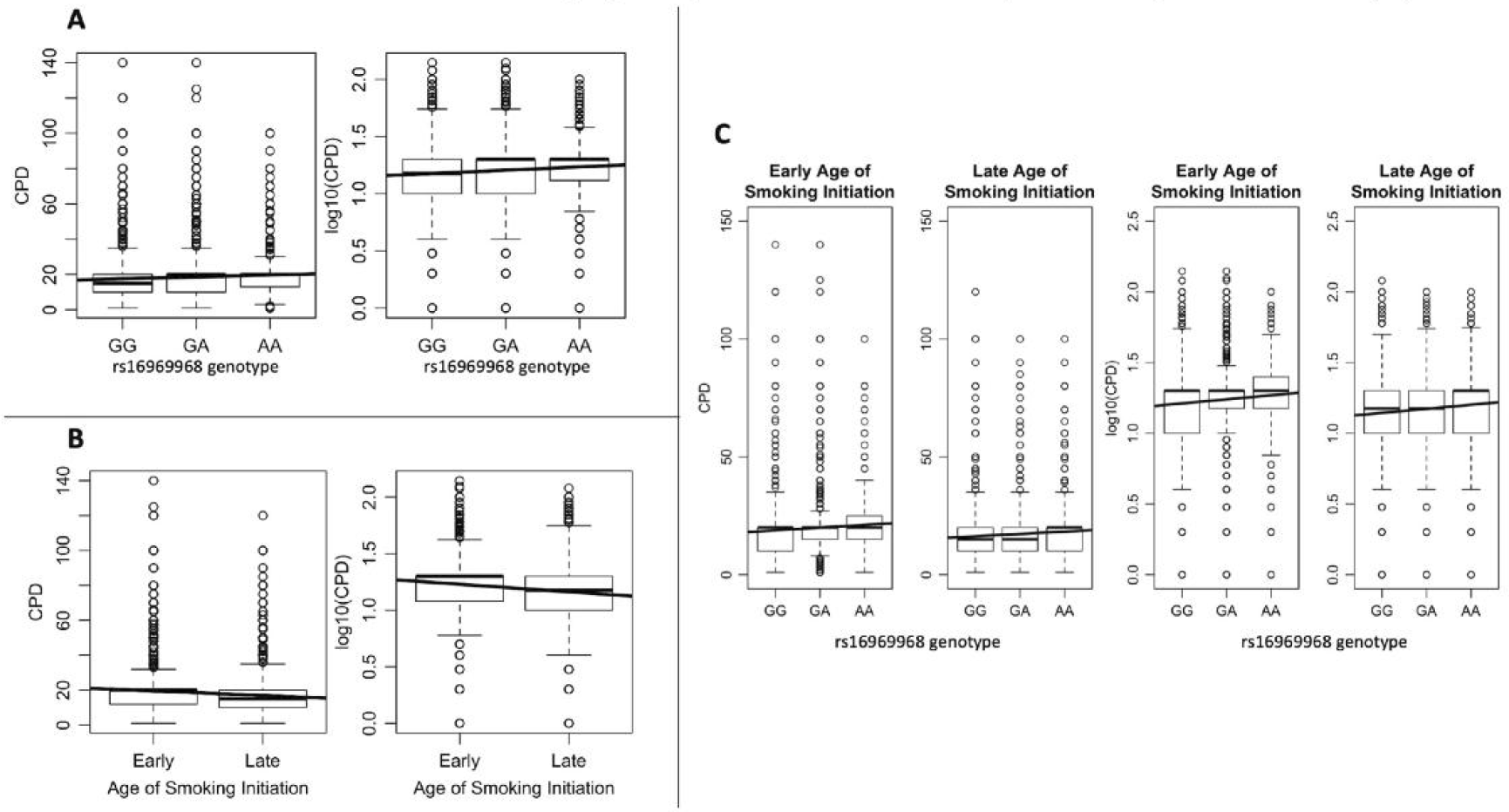
Main effects of rs16969968 genotype (A) and AOS (B) on cigarettes per day (CPD or log_10_(CPD)), and rs16969968 effect on CPD and log_10_(CPD) as a function of early or late age of initiation (C).

Conversely, the interaction between rs16969968 genotype and AOS was only nominally significant (α=0.05) and only in some combinations of CPD and AOS encodings (Figures 1C, S4; Tables 1, S1-2). Specifically, when treating both CPD and AOS as binary phenotypes the logistic model interaction was not significant (ORrs16969968×AOS=1.004, *p*=0.82) and the effect was notably lower than the previously reported estimate of 1.16. Interestingly, the interaction effect was nominally significant (p<0.05) for the binned CPD phenotype and dichotomized AOS, and when heavy/light CPD was analyzed on the additive scale, but not when the CPD phenotype was either heavy vs. light analyzed on the multiplicative scale model or when CPD was log-transformed. Across all tests and all CPD and AOS encodings, no interaction effects reached genome-wide significance (*p*>0.028).

Associations of rs16969968 stratified by AOS also produced mixed results. 95% confidence intervals (α=0.05) of the meta-analyzed effect sizes were non-overlapping only for binned and binary CPD encodings (Fig. 2, Table S3). When examining meta-analyzed genetic effects of rs16969968 on heavy versus light CPD, OREarly/ORLate was much lower than previously reported (OREarly/ORLate=1.016, Table S3). Within the largest ancestry cluster (EUR-ancestry), 95% CIs of the rs16969968 effects were non-overlapping in early vs. late AOS individuals for all CPD encodings except log_10_(CPD) (Fig S5; Table S3-S5). For all other ancestry clusters, we found no evidence of different rs16969968 effects using either a genome-wide (α=5e-8) or nominal (α=0.05) significance threshold (Table S4).

**Figure 2.**
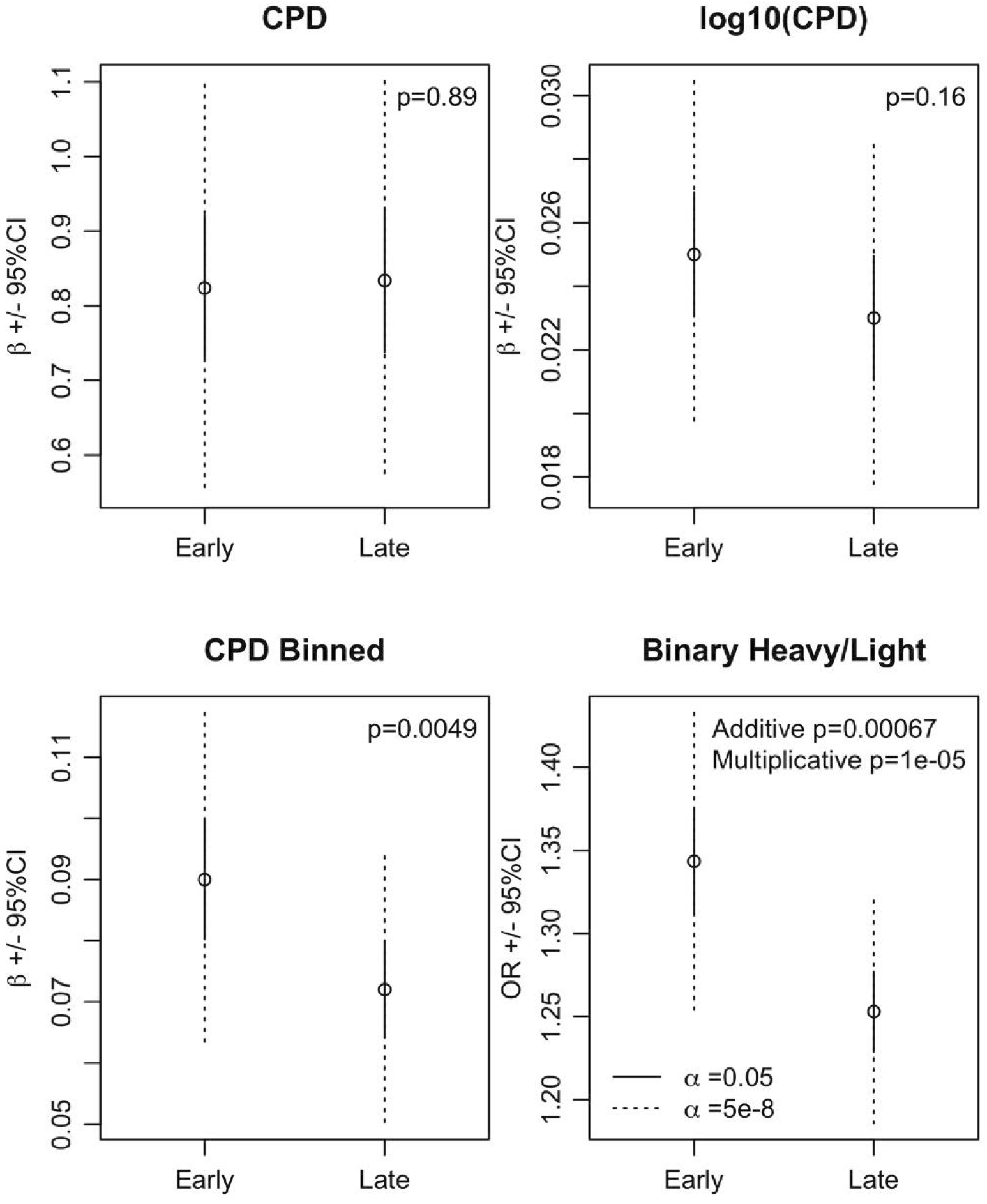
Trans-ethnic meta-analyzed allelic effect size estimates, *β*, of the rs16969968 risk allele, A, estimated in stratified association analyses by early and late age of initiation, with CIs indicated using either *α*=0.05 (solid line) or genome-wide Bonferroni-corrected *α*=5e-8 (dashed line). For the heavy vs. light smoker phenotype, allelic effects (*β*) were transformed to OR using the BOLT-LMM^38^- suggested transformation, of e*^β^*^/(0(1-^*^u^*^))^, where *u=*0.42 is the proportion of cases. Effect size difference Z-test p-values are shown for each comparison. See Table S3 for estimates and statistics.

The direct replication test using Hartz et al.^13^ methods with only rs16969968 and sex as independent variables found no evidence of different allelic effects between early and late smokers (p=0.41; Table S6-S8).

Our power analyses yielded two main results. First, our sample was well powered (>99%) to detect an interaction effect of the size previously reported at nominal significance (Figs. 3, S1), though not at genome-wide significance (power ~5%), even with over 61,000 subjects. Second, a sample drastically larger than that analyzed here would be required to detect an interaction effect of ORrs16969968×AOS=1.004, as estimated within our sample, with 80% power at α=0.05 (Fig. 3). Even applying the upper 95% CI limit of our estimate (ORrs16969968×AOS=1.035), or the estimate within the largest genetic cluster (1.03) would require a sample of approximately 1.3-2 million participants to achieve 80% power at a genome-wide threshold (α=5e-8) (Figs. S6, S7).

**Figure 3.**
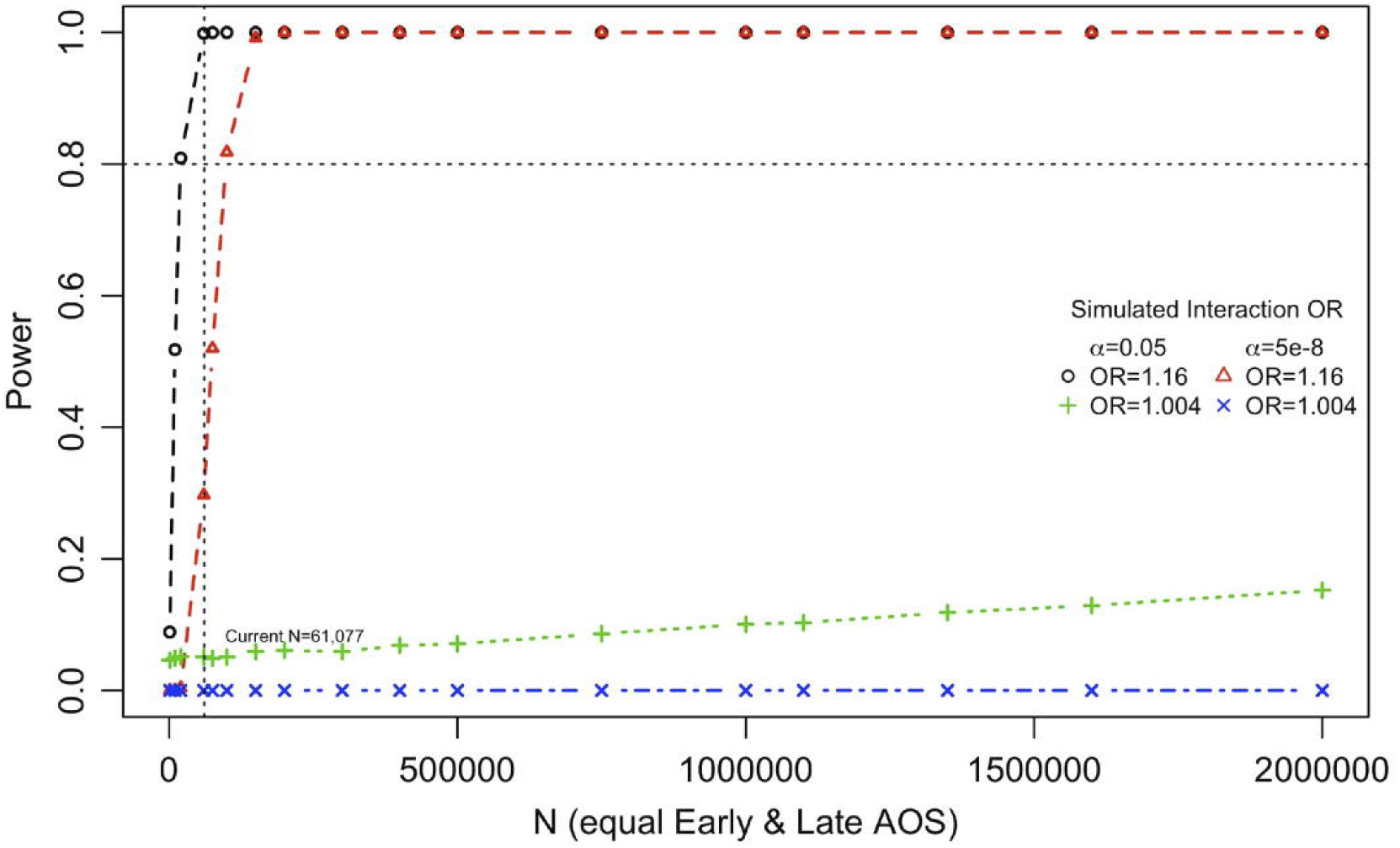
Power to detect the statistical interaction effect across a range of sample sizes for the interaction effect size estimated previously (OR=1.16) and from the current study (OR=1.004), applying either a nominal *α*=0.05 or genome-wide Bonferroni-corrected *α*=5e-8.

## DISCUSSION

We replicated the substantial main effects of rs16969968 and early age of onset of smoking on CPD, across all phenotypic and analytical scales (Table 1). Estimates were in the same direction and of roughly similar magnitude as those previously reported^13^.

Conversely, we found limited evidence of an rs16969968×AOS interaction effect. Formal interaction model results were mixed and depended heavily on measurement scale and phenotype encoding. Notably, our attempt to directly replicate the methods of Hartz et al. failed to identify a significant difference in the rs16969968-A allele effect on heavy smoking between early and late AOS (p=0.41; Table S6). This is similar to the results from stratified linear mixed model analyses, where the genetic effect in early AOS individuals was 1.016-fold higher than in late AOS individuals, despite greater statistical power of linear mixed models^39^, as well as more control of potential confounding variables, such as genetic ancestry and geographic variation throughout the UK. Patterns within the largest genetic ancestry cluster, individuals with primarily EUR ancestry, were similar to the trans-ethnic meta-analyzed results. While some tests of differences in the stratified associations did reach nominal significance, the results suggest only minute differences in rs16969968 effects between early and late initiating smokers (Table S3). Across multiple analytic frameworks and phenotype encodings, the majority of our results were incongruent with an interaction between rs16969968 and AOS.

### Magnitude of Effects and Power

No interaction test, and no comparison of stratified estimates, reached genome-wide significance (α=5e-8) despite the comparatively large sample size of our study. With genome-wide genotyping arrays and imputation commonly applied^41^, and as genome-wide interaction associations and heritability studies have become more frequent^42-48^, focusing on genome-wide significance thresholds is paramount to avoid false positives, even in situations where there are *a priori* hypotheses of interaction, as in rs16969968×AOS. Applying sufficiently stringent significance thresholds in initial studies, whether genome-wide 5e-8 or another specified threshold, is a best practice for replication of association studies^49^, and we believe that as GWAS interactions (including G×E and G×G) studies become more frequent, the question of applicable significance thresholds should be revisited.

In all tests related to interaction effects and stratified associations, the estimated interaction effect sizes were much smaller than previously reported^13^. Despite substantially greater power than the original study, which had a sample size of ~30,000, (Fig. 4, S6-S7), we estimated the effect to be only 1.004 (or 1.016 in the stratified associations). The lack of replication when using the exact same methods suggests that there is no true interaction at this locus. It is important to recognize that both replicated main effects were strong, significant, and in the expected direction, reflecting the strongest single-locus genetic effect on CPD^12^ and a strong, consistent risk factor of heavy smoking (early age of initiation). This suggests that if the interaction were to exist, its effect would be much less than previously expected. Importantly, with an OR=1.004, it would be insignificant for possible clinical interventions, such as targeted smoking awareness based on rs16969968 genotype^26^.

The discrepancy between our results and those reported by Hartz et al.^13^ could additionally reflect differences between the study populations and models used for analyses. The study by Hartz et al.^13^ exemplified a tremendous effort to collect the largest available sample size at the time. They were able to do so by meta-analyzing multiple individual studies together, a highly coordinated endeavor that must be recognized and applauded. One possible outcome of this approach is heterogeneity of effect estimates, which they found and noted. Our analysis focused on a single, relatively homogeneous dataset instead of many studies, removing potential heterogeneity that could have influenced the previous results. Our meta-analysis of relatively homogenous ancestry clusters also attempted to minimize any confounding of stratification. Second, although 33% of the Hartz et al. data were European datasets, consistent cultural differences may exist between American and UK samples, such as general attitudes towards smoking, and any potential impact would be difficult to assess. Such differences between samples could lead to true heterogeneity in the effects^50^ and the different estimated effects we observed, though Hartz et al. reported no significant difference between OR estimates from American versus EU studies. A possible source of bias, in both the initial and the current study, is that of collider bias^51,52^. The UK Biobank is healthier and wealthier^51^ than the general UK Population, leading to ascertainment and the potential for colliders. Genetic effects on education could lead to false negative genetic associations in the UK Biobank with smoking traits when controlling for education^51^, but we view this as an unlikely explanation for our failure to replicate results, as both main effects were replicated, and because whether we controlled for education or not, we found little evidence of rs16969968×AOS interaction. Selection bias in general could lead to false positive or negative associations. Additional methodological differences include testing a full statistical interaction model with complete covariate×AOS and covariate×genotype terms and using a linear mixed model in our stratified analyses, neither of which were previously employed. Mixed model approaches generally improve power^39^, and including the covariate interaction terms should lead to unbiased estimates of the rs16969968×AOS interaction^37^. On the other hand, comparing estimates across different subsamples, as in stratified linear mixed model analysis, introduces an additional potential source of confounding. However, the respective strengths and weaknesses of these methods cannot account for our failure to directly replicate the original finding; our stratified association tests with only sex as a covariate (mirroring the approach of Hartz et al.) failed to identify significant differences in allelic effect sizes between early and late AOS individuals (p=0.41; Table S6), despite being well-powered to do so.

Regardless, with respect to particular phenotype encodings and analyses (e.g., stratified analyses of heavy vs. light smoker status, with linear mixed models), we did find nominally significant, very small differences in allelic effect size estimates between early- and late-onset smokers. These findings are thus potentially congruent with a small interaction between rs1696968 and AOS. If there is a true rs16969968×AOS interaction of roughly the magnitude we estimated (OR=1.004), it would a drastically larger sample size to detect it (Fig. 4). We must therefore conclude that any such interactions specific to an individual locus are likely of very small effect, will be very difficult to identify even with the largest available biobanks, and likely contribute minimally to phenotypic variance.

### Conclusions

We found limited support for the rs16969968×AOS interaction. To the extent that AOS might moderate the effect of rs16969968, we estimate this effect to be far smaller than previously reported. We suggest that even larger sample sizes will be required to identify, with genome-wide significance, interactions at individual loci given the expected magnitude of the interaction effects. On the other hand, our unambiguous replications of the main effects of both rs16969968 and AOS on smoking quantity support epidemiological evidence that individuals who begin regularly smoking at a young age are at a higher risk for nicotine dependence later in life^21,22^. This provides further evidence in support of public health interventions for adolescent smoking that could help reduce tobacco use, which would in turn lower the number of tobacco-related deaths and illnesses.

## Data Availability

The data used are available through the UK Biobank.

https://www.ukbiobank.ac.uk/

## ACKNOWLEDGEMENTS

Ms. Adjangba was supported by the Summer Multicultural Access to Research Training Program at the University of Colorado. Drs. Evans and Border are supported by National Institute of Mental Health R01 MH100141-06 (PI: Matthew C. Keller) and Dr. Evans is supported by National Institute on Drug Abuse R01 DA044283-01A1 (PI: Scott I. Vrieze) and National Institute on Aging R01 AG046938 (PI: C.A. Reynolds/S.M. Wadsworth).

## Conflict of Interest Disclosures

The authors declare no conflict of interest.

## Notes

### Competing Interest Statement

The authors have declared no competing interest.

### Clinical Protocols

https://osf.io/ynh2j

### Summary of Updates

Clarification of the importance of validation and replication of association tests. Utilizing the entire UK Biobank sample across all ancestries.

